# Dynamic Graph Transformer for Brain Disorder Diagnosis

**DOI:** 10.1101/2024.09.05.24313048

**Authors:** Ahsan Shehzad, Dongyu Zhang, Shuo Yu, Shagufta Abid, Feng Xia

## Abstract

Dynamic brain networks are crucial for diagnosing brain disorders, as they reveal changes in brain activity and connectivity over time. Previous methods exploit the sliding window approach on fMRI data to construct these networks. However, this approach encounters two major issues: fixed temporal length, which inadequately captures the temporal dynamics of brain activity, and global spatial scope, which introduces noise and reduces sensitivity to localized dysfunctions when applied across the entire brain. These issues can lead to inaccurate brain network representations, potentially resulting in misdiagnosis. To overcome these challenges, we propose BrainDGT, a dynamic Graph Transformer model that adaptively captures and analyzes modular brain activities for improved diagnosis of brain disorders. BrainDGT addresses the fixed temporal length issue by estimating adaptive brain states through deconvolution of the Hemodynamic Response Function (HRF), avoiding the constraints of fixed-size windows. It also addresses the global spatial scope issue by segmenting fMRI scans into functional modules based on established brain networks for detailed, module-specific analysis. The model employs a dual attention mechanism: graph-attention extracts structural features from dynamic brain network snapshots, while self-attention identifies significant temporal dependencies. These spatio-temporal features are adaptively fused into a unified representation for disorder classification. BrainDGT’s effectiveness is validated through classification experiments on three real fMRI datasets ADNI, PPMI, and ABIDE demonstrating superior performance compared to state-of-the-art methods. BrainDGT improves brain disorder diagnosis by offering adaptive, localized analysis of dynamic brain networks, advancing neuroimaging and enabling more precise treatments in biomedical research.

## I. Introduction

**T**HE diagnosis of brain disorders via brain network analysis is a rapidly evolving research area, promising significant advancements in early disorder detection and intervention [1]. Functional Magnetic Resonance Imaging (fMRI) is pivotal in this domain, modeling brain networks by measuring neural activity [2]. fMRI detects blood flow changes associated with neural activity, enabling detailed mapping of functional brain connectivity. This method is crucial for diagnosing brain disorders, as it can reveal network connectivity abnormalities undetectable by traditional imaging techniques. Analyzing these networks helps clinicians identify patterns linked to specific neurological disorders, leading to earlier and more accurate diagnoses [3]. Brain network analysis’s potential lies in uncovering subtle brain function changes, providing a powerful tool for understanding and diagnosing complex brain disorders.

Over the years, various methods have been developed for brain network analysis to diagnose brain disorders [4], [5]. These methods can be divided into static and dynamic brain network analysis methods [6]. Static brain networks are commonly constructed from fMRI data using the Pearson correlation method, which measures the connectivity between different brain regions based on the correlation of their activity over time [2], [7]. Traditionally, these networks are analyzed using graph-based structural features combined with machine learning algorithms or end-to-end graph learning methods [8]. However, static methods are inadequate for capturing the timevarying changes in brain connectivity that are crucial for understanding the progression of brain disorders. In contrast, dynamic brain networks are constructed from fMRI data using the sliding window method, which captures temporal fluctuations in brain connectivity [9]. These networks are analyzed using spatio-temporal graph learning techniques [10]. Despite their advantages, they face limitations such as a restricted receptive field, which can hinder their ability to fully capture complex brain dynamics [11]. Recent advancements in graph learning have introduced Graph Transformers, which are powerful tools for capturing complex dependencies in graph-structured data [12]. Graph Transformers have shown significant potential in various applications due to their ability to model long-range interactions and contextual information [13], [14]. In the context of brain network analysis, a few Graph Transformer-based methods have been applied to dynamic brain networks for diagnosing brain disorders, demonstrating improved performance over traditional methods by better capturing the intricate temporal dynamics of brain connectivity [15], [16].

However, previous methods of dynamic brain network analysis for diagnosing brain disorders often rely on the sliding window approach to construct dynamic brain networks, which has two major issues. The first issue is the use of fixed temporal length windows. These fixed-length windows fail to capture the temporal dynamics of brain activity adequately (see Fig.1). Brain activity fluctuates over time, and a rigid window length cannot adapt to these fluctuations, resulting in an incomplete or distorted representation of brain activity [17]. The second issue is the global spatial scope of these methods. They apply full-width windows across all Blood-Oxygen-Level Dependent (BOLD) signals of the entire brain, rather than focusing on individual functional modules. This broad approach introduces noise and diminishes sensitivity to localized dysfunctions [18]. Consequently, these issues lead to inaccurate brain network representations. Inaccurate representations can significantly impact the diagnosis process, potentially resulting in misdiagnosis and inappropriate treatment plans for patients with brain disorders.

**Fig. 1.**
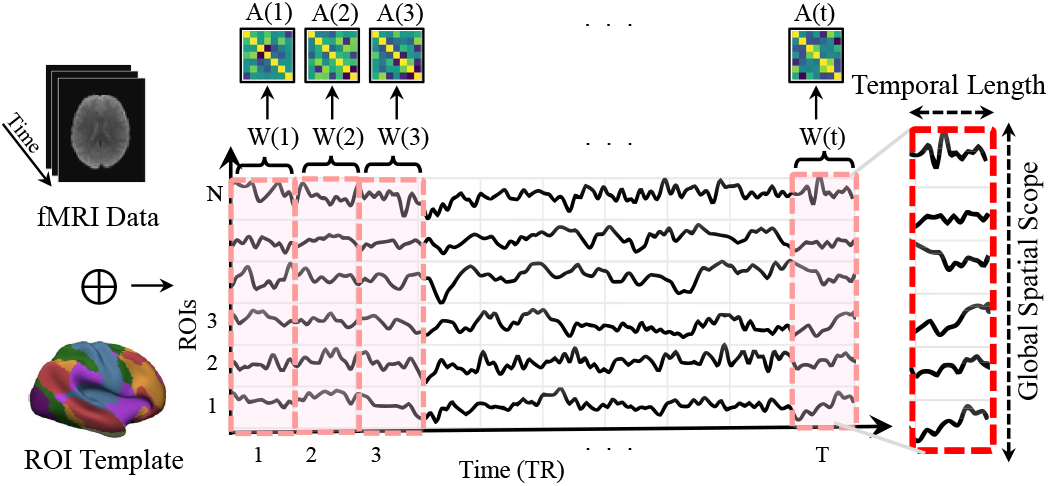
Conventional sliding window based dynamic brain networks suffer with two major issues: (1) Fixed Temporal Length: Inadequate capture of brain activity dynamics. (2) Global Spatial Scope: Full-width windows introduce noise and reduce sensitivity to localized dysfunction, leading to fully connected networks that challenge traditional Graph Transformers.

To address these challenging issues, we introduce BrainDGT, a unified framework for enhancing dynamic brain network construction and disorder diagnosis. It addresses the fixed temporal length issue by estimating adaptive brain states via deconvolution of the Hemodynamic Response Function (HRF), capturing the variable nature of brain activity more accurately. To address the problem of global spatial scope, BrainDGT segments fMRI scans into functional modules based on established brain networks, allowing detailed module-specific analysis, reducing noise, and increasing sensitivity to localized dysfunctions. BrainDGT also employs dynamic Graph Transformers with a dual attention mechanism: graph-attention learns structural features from each network snapshot, while self-attention captures temporal dependencies. These spatio-temporal features are fused for classification (diagnosis), ensuring comprehensive analysis. We validated BrainDGT on three fMRI datasets (ADNI, PPMI, and ABIDE). BrainDGT demonstrates superiority over state-of-the-art methods, highlighting its potential for improved detection of brain disorders. Key innovations and contributions include:

1. We propose BrainDGT, a dynamic brain Graph Transformer model that enhances brain disorder diagnosis by adaptively capturing the temporal dynamics of brain connectivity within individual functional brain modules, thereby addressing the issues of fixed temporal length and global spatial scope.
2. BrainDGT segments fMRI scans into functional modules based on established brain networks for detailed analysis. It estimates adaptive brain states within each module via deconvolution of the HRF, capturing brain activity’s temporal dynamics. BrainDGT uses dynamic Graph Transformers with dual attention to extract and fuse key spatio-temporal features for effective brain disorder detection.
3. We validate the effectiveness of BrainDGT through extensive classification experiments on three real fMRI datasets: ADNI, PPMI, and ABIDE. Experimental results demonstrate that BrainDGT outperforms state-of-the-art methods.

The remainder of this paper is structured as follows: Section II reviews related work, highlighting key contributions and identifying gaps that this study aims to address. Section III describes the design of BrainGT, emphasizing its architecture and implementation. Section IV presents and discusses the experimental results, evaluating the performance and implications of the proposed design. Finally, Section V concludes the paper and suggests directions for future research.

## II Related Work

### A. Brain Network Analysis

Brain network analysis is crucial for diagnosing brain disorders by examining structural and functional connectivity to identify biomarkers and patterns associated with neurological conditions [19], [20]. These networks are categorized into static and dynamic types, each with distinct assumptions, construction methods, and analytical approaches [6]. Static brain networks assume stable connectivity during the scan, offering a single, unchanging snapshot of brain activity [18], [21]. Typically, these networks are built using the Pearson correlation method, which quantifies the linear relationship between time series of different brain regions of interest (ROIs) [22]. Traditional machine learning techniques extract graph theoretical features from the connectivity matrix, such as node degree, clustering coefficient, and path length, to classify healthy and diseased states [19], [20]. Recent advancements include graph learning techniques, where models are trained end-to-end on brain connectivity data. For example, BrainGNN uses graph neural networks to analyze fMRI data, featuring ROI-aware graph convolutional layers and ROI-selection pooling layers [23]. BrainGB provides a standardized platform for evaluating GNN models on brain network datasets, supporting various architectures for neuroscience research [4]. BRAIN-NETTF employs Transformer-based architecture to model brain networks as graphs and identify functional brain modules using an orthonormal clustering readout [24]. However, static brain networks fail to capture time-varying changes, offering only a single snapshot and potentially overlooking dynamic interactions and transient states critical for understanding brain disorders [25]. Consequently, static analyses may miss nuanced connectivity patterns over time, limiting their diagnostic power [26].

Dynamic brain networks assume that brain connectivity varies over time, reflecting the brain’s dynamic nature [27]. This approach considers temporal fluctuations in connectivity, capturing the brain’s adaptive and transient states [28]. The sliding window method segments the fMRI time series into overlapping or non-overlapping windows, calculating connectivity matrices for each window. This creates time-resolved connectivity matrices that show the evolution of brain connectivity. Gadgil et al. [29] presented the ST-GCN model to analyze resting-state fMRI data by capturing dynamic functional connectivity as spatio-temporal graphs. Dahan et al. [30] introduced the ST-fMRI model, leveraging spatio-temporal dynamics with graph convolutional networks and multi-resolution ICA nodes. Kim et al. [15] developed STAGIN, a model focusing on spatio-temporal attention to learn dynamic graph representations using novel READOUT functions and a Transformer encoder. Despite their promise, fixed window lengths in many studies fail to capture the full temporal dynamics of brain activity. The assumption of constant window size does not account for varying durations of different brain states, potentially missing critical transitions and interactions. Consequently, while dynamic brain networks offer a more comprehensive view, their construction and analysis methods need refinement to fully capture the brain’s temporal complexity.

### B. Dynamic Graph Transformers

Dynamic Graph Transformers (DGTs) integrate graph neural networks with Transformer architectures to capture spatial and temporal dependencies in graph-structured data [31]. Using self-attention mechanisms, DGTs effectively model intricate and dynamic relationships within graph data, making them suitable for applications with changing node interactions [32]. These models are applied in social network analysis [33], transportation networks [34], and neuroscience [35], particularly for diagnosing and analyzing brain disorders by capturing the temporal evolution of brain connectivity [36].

Fortunately, Some studies have demonstrated the potential of DGTs in brain disorder diagnosis. Kan et al. [16] proposed DART, integrating static and dynamic brain networks using a Graph Transformer with a Multi-Head Self-Attention Module. Ma et al. [37] introduced MDGL, constructing multi-scale dynamic functional connectivity networks from fMRI time series and employing dynamic graph representation learning. Zhu et al. [38] developed OT-MCSTGCN, using optimal transport theory to track topology evolution in dynamic brain networks. However, existing methods fail to adaptively capture dynamic brain activity within specific functional modules. BrainDGT addresses these limitations by constructing individual dynamic brain networks for each functional module, identifying adaptive brain states through HRF deconvolution, and capturing modular brain activities to improve disorder detection accuracy.

## III. The Design of BrainDGT

### A. The Overview of BrainDGT

BrainDGT is a dynamic brain Graph Transformer model designed to enhance the diagnosis of brain disorders by adaptively capturing and analyzing modular brain activities. The framework begins by dividing fMRI scans into functional modules based on established brain functional networks. Within each module, brain activity is captured using the deconvolution of HRF, which represents temporal sequences of brain states. Dynamic functional connectivity is then calculated for each brain state to construct a dynamic brain network for each functional module. Self-attention-based graph encoders learn structural features from each network snapshot, while Transformer-based temporal encoders extract temporal dependencies between snapshots. The spatio-temporal features from all temporal encoders are adaptively integrated into a unified representation, which is then processed through a Multi-Layer Perceptron (MLP) for disorder classification. The framework and workflow of BrainDGT are depicted in Fig 2.

**Fig. 2.**
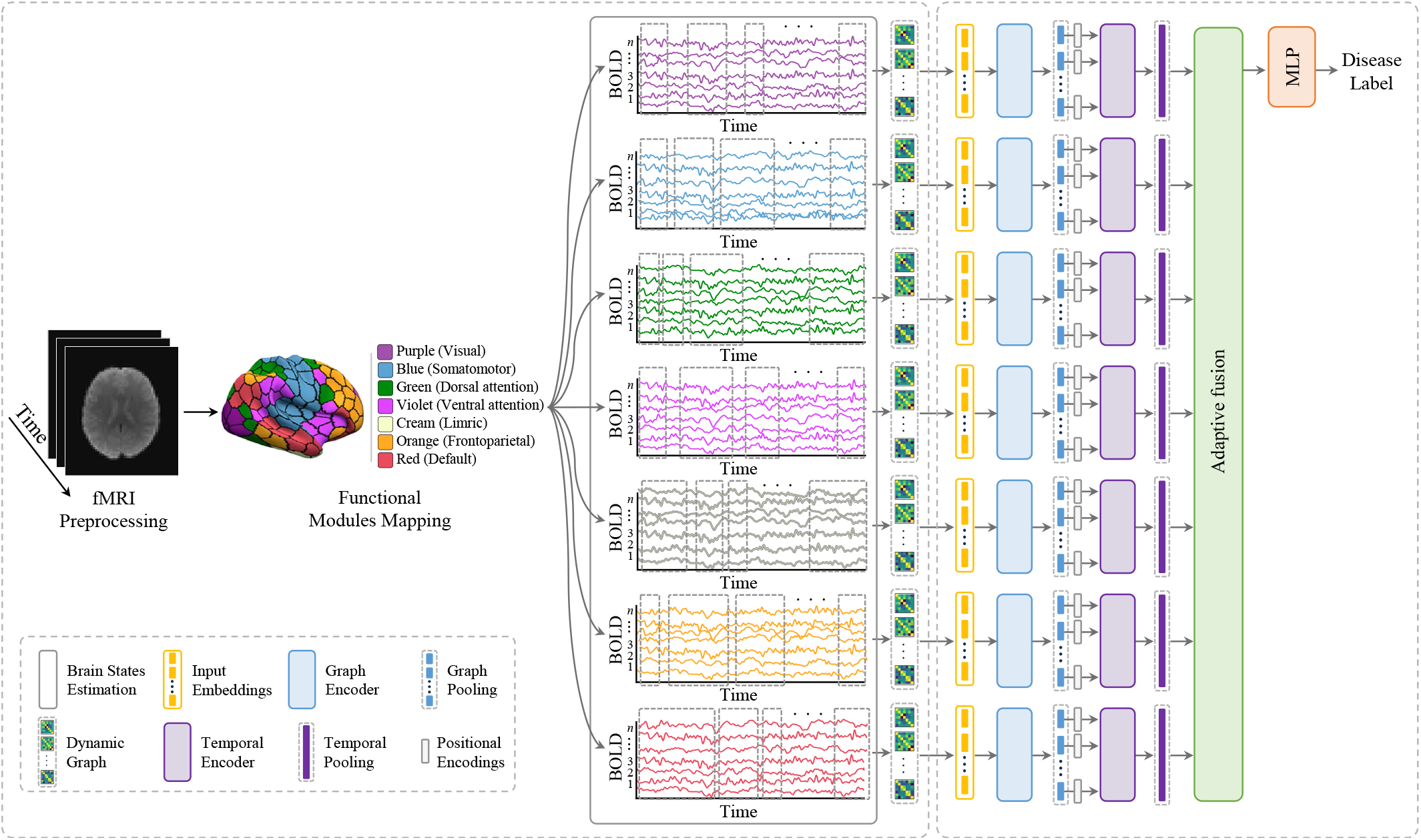
The framework of BrainDGT.

### B. Dynamic Brain Network Construction

#### 1) Data Preprocessing

Preprocessing functional magnetic resonance imaging (fMRI) data is a crucial step in our neuroimaging research, ensuring that the raw data is refined and ready for subsequent analysis [18]. This process involves several key steps: slice timing correction, motion correction, normalization to a standard space, and spatial smoothing. Each step addresses specific issues inherent in fMRI data acquisition and enhances the quality and interpretability of the resulting data.

##### a) Slice Timing Correction

Slice timing correction addresses the temporal discrepancies in data acquisition across different slices of the brain. During fMRI scanning, slices are captured sequentially, not simultaneously, which leads to temporal offsets that can introduce artifacts, especially in fast-changing brain activities. To correct for these offsets, we interpolate each voxel’s time series to match a reference time point, often the acquisition time of the middle slice. If *T*_*ijk*_(*t*) represents the signal intensity at voxel (*i, j, k*) at time *t*, and Δ*t* is the time difference between slices, we resample *T*_*ijk*_(*t*) to a common temporal reference using interpolation methods such as cubic spline interpolation or Fourier-based interpolation:

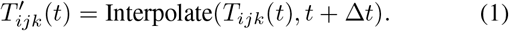

##### b) Motion Correction

Motion correction compensates for head movements that occur during the scanning session, which can cause significant artifacts and misinterpretations in the data. This step involves realigning all volumes in the time series to a reference volume, typically the first or middle volume. We achieve this through rigid-body transformations, which include translations and rotations. The objective function we minimize during motion correction is often the sum of squared differences (SSD) or correlation ratio (CR) between the reference volume and subsequent volumes. The transformation parameters *θ* = (*t*_*x*_, *t*_*y*_, *t*_*z*_, *α, β, γ*) (translations and rotations along each axis) are optimized such that:

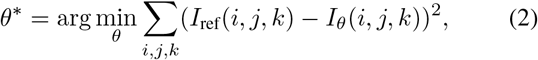

where *I*_ref_ is the intensity of the reference volume and *I*_*θ*_ is the transformed intensity.

##### c) Normalization

Normalization involves transforming individual brain images into a common stereotactic space, such as the Montreal Neurological Institute (MNI) space. This step allows for inter-subject comparisons and group analysis by aligning anatomical structures across different subjects. Normalization typically involves a two-step process: linear transformation (affine registration) followed by non-linear warping. The affine transformation aligns the images roughly by scaling, rotating, and translating them to match a standard template. Non-linear warping then adjusts for finer anatomical differences. We represent the transformation as:

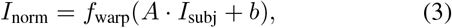

where *A* is the affine transformation matrix, *b* is the translation vector, and *f*_warp_ represents the non-linear deformation field.

##### d) Spatial Smoothing

Spatial smoothing is applied to reduce high-frequency noise and improve the signal-to-noise ratio (SNR). It also helps accommodate anatomical variability across subjects by blurring the data to a certain extent. This step involves convolving the fMRI data with a Gaussian kernel *G*_*σ*_(*x, y, z*) characterized by its full-width at half-maximum (FWHM), which determines the extent of smoothing. The smoothed signal 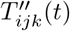 at voxel (*i, j, k*) and time *t* is given by:

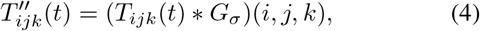

where∗ denotes the convolution operation. Our choice of *σ* (typically 4-8 mm FWHM) balances the trade-off between noise reduction and spatial resolution.

#### 2) Functional Module Mapping

The process of mapping fMRI scans into functional modules involves dividing the brain into distinct regions based on functional connectivity patterns. In our study, we employ the Schaefer Atlas^1^, particularly the version that partitions the brain into 400 parcels [39]. This granularity strikes a balance between detailed spatial resolution and computational manageability. To apply the Schaefer Atlas to preprocessed fMRI data, we first ensure that the data is registered to a standard anatomical space, such as the MNI space. This registration aligns the spatial coordinates of the Schaefer Atlas with the fMRI data. Next, we assign each voxel in the preprocessed fMRI data to one of the 400 parcels defined by the Schaefer Atlas. This is achieved by overlaying the atlas on the fMRI data and labeling each voxel according to the parcel it falls into. Mathematically, if *V* represents the set of voxels and *P*_*i*_ denotes the *i*-th parcel, then each voxel *v∈ V* is assigned a label *l*_*v*_ where *l*_*v*_ = *i* if *v∈ P*_*i*_, with *i* ranging from 1 to 400.

The Schaefer Atlas also provides a mapping of each parcel to one of seven predefined functional networks: Visual, Somatomotor, Dorsal Attention, Ventral Attention, Limbic, Frontoparietal, and Default Mode. We use a majority voting algorithm to assign each of the 400 parcels to these networks based on voxel overlap with the predefined networks. For each parcel *P*_*i*_, we determine the number of voxels within *P*_*i*_ that overlap with each network *N*_*j*_. We then assign parcel *P*_*i*_ to the network *N*_*j*∗_ for which the overlap is maximized, where

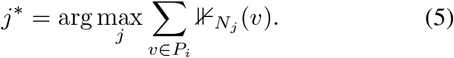

Here, ⊮_*Nj*_ (*v*) is an indicator function that equals 1 if voxel *v* belongs to network *N*_*j*_, and 0 otherwise. The final network assignment for each parcel is determined by Network(*P*_*i*_) =*N*_*j*∗_.

#### 3) Brain State Estimation using HRF

Estimating brain states from fMRI data involves modeling the underlying neural activity that gives rise to observed blood-oxygen-level-dependent (BOLD) signals. The HRF characterizes the relationship between neural activity and BOLD signals [40]. The first step in estimating brain states is extracting the BOLD signals from the preprocessed fMRI data. This involves selecting functional modules and averaging the BOLD signals within each module to obtain a representative time series. Let *T*_*Nj*_ (*t*) represent the BOLD signal at time *t* for functional module *N*_*j*_:

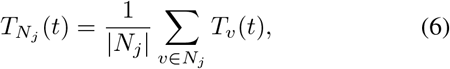

where *T*_*v*_(*t*) is the BOLD signal at voxel *v*, and |*N*_*j*_| is the number of voxels in module *N*_*j*_.

The HRF is typically modeled using a double gamma function, which captures the primary and secondary peaks of the hemodynamic response [41]. The double gamma function *h*(*t*) is defined as:

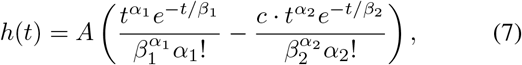

where *α*_1_, *β*_1_, *α*_2_, *β*_2_ are parameters defining the shape of the function, *A* is a scaling factor, and *c* is the ratio of the second gamma function’s amplitude to the first.

To estimate neural activity *n*(*t*) from the observed BOLD signal *T* (*t*), we use Wiener deconvolution, a technique to reverse the convolution of the neural activity with the HRF. The observed BOLD signal can be expressed as:

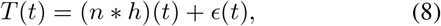

where ∗ denotes convolution and *ϵ*(*t*) represents noise. The Wiener deconvolution filter [42] *H*_*w*_(*f*) in the frequency domain is given by:

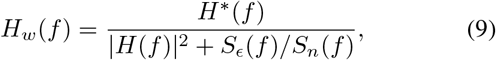

where *H*(*f*) is the Fourier transform of the HRF, *H*^∗^(*f*) is its complex conjugate, *S*_*ϵ*_(*f*) is the power spectral density of the noise, and *S*_*n*_(*f*) is the power spectral density of the neural activity. The estimated neural activity 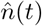 is obtained by:

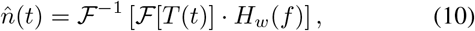

where *ℱ*and *ℱ*^−1^ denote the Fourier and inverse Fourier transforms, respectively.

Change point detection is used to identify peaks in the estimated neural activity, which correspond to transitions between distinct brain states. This involves detecting significant changes in the time series that indicate shifts in neural activity patterns. Let 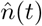 be the estimated neural activity. Change point detection method, the Pruned Exact Linear Time (PELT) algorithm [43], are applied to 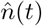 to identify time points *{t*_1_, *t*_2_, …, *t*_*k*_*}* where significant changes occur:

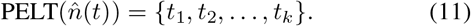

Once the change points are identified, the time series is segmented into intervals representing distinct brain states. Each segment *S*_*i*_ between change points *t*_*i*_ and *t*_*i*+1_ is considered a distinct brain state:

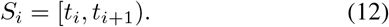

These segments provide a temporal representation of brain states, which can be further analyzed to understand the dynamics of brain activity. Finally, we construct a sequence of brain states for each functional ROI based on identified events *S*_*j =*_ *{s*_1_, *s*_2_, …, *s*_*k*_*}*, where each *s*_*k*_ represents a neural event segment. The conceptual representation of brain states estimation is given in Fig. 3.

**Fig. 3.**
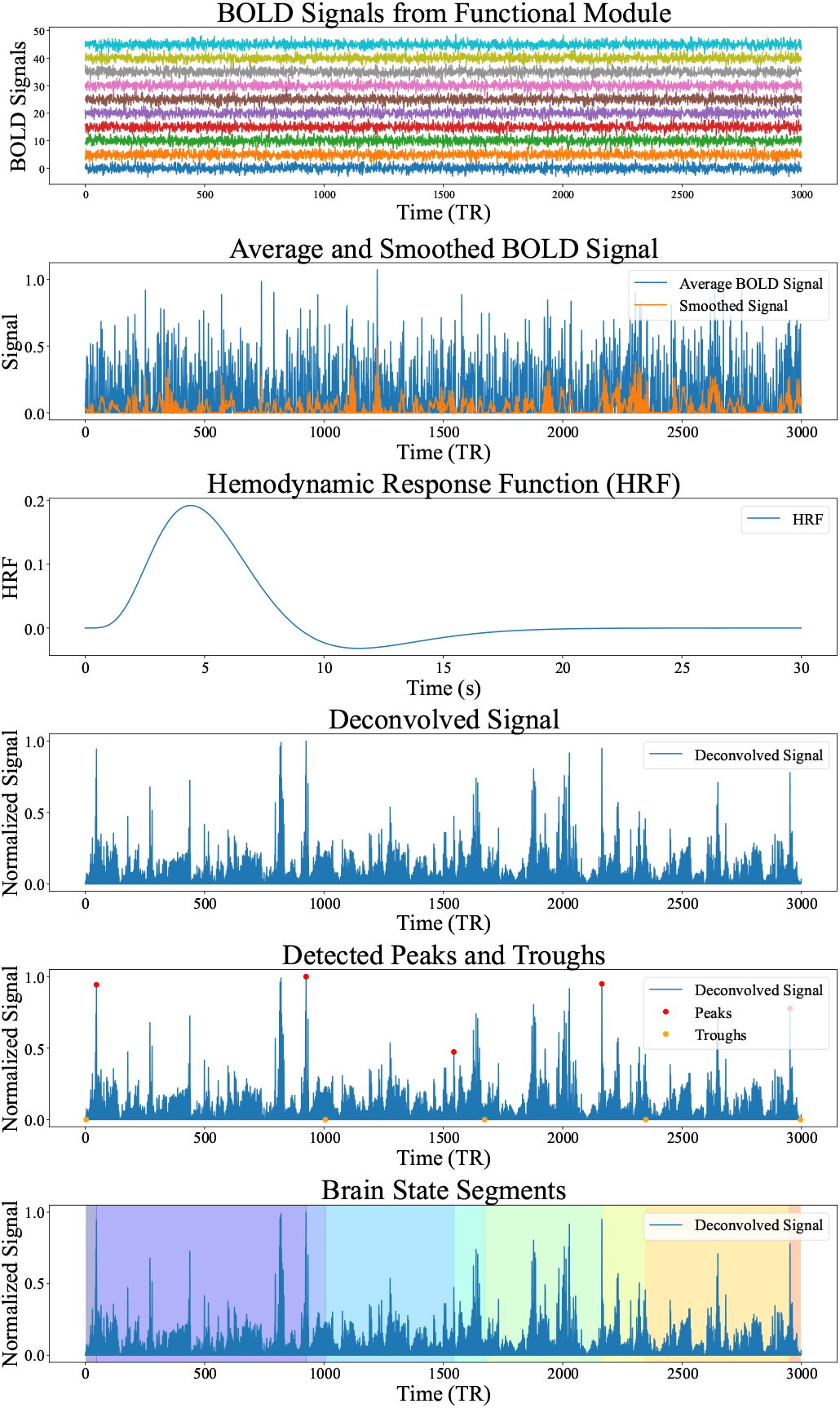
Estimation of brain staes using deconvolution of HRF.

#### 4) Dynamic Connectivity Estimation

Dynamic connectivity estimation involves calculating time-varying functional connectivity metrics between brain regions to understand the changing interactions within the brain over time. Functional connectivity between brain regions is typically measured using metrics such as Pearson correlation. For each brain state, represented by the segmented BOLD signals, we calculate th, e Pears(o7n) correlation coefficient between the time series of different brain regions. Let 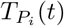 and 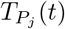 represent the BOLD signal time series for parcels *P*_*i*_ and *P*_*j*_ during a brain state *S*. The Pearson correlation coefficient *r*_*ij*_ is computed as:

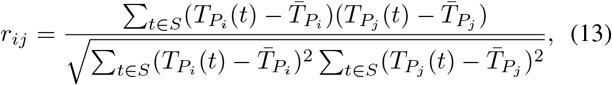

where 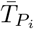*i* and 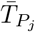 are the mean BOLD signals of parcels *P*_*i*_ and *P*_*j*_over the state *S*. For each brain state, the functional connectivity metrics are used to construct a dynamic brain network. A dynamic brain network is a graph where vertices represent brain regions (parcels), and edges represent the functional connectivity between them. Each parcel *P*_*i*_ is represented as a vertex *v*_*i*_ in the graph, and each edge *e*_*ij*_ between vertices *v*_*i*_ and *v*_*j*_ is weighted by the Pearson correlation coefficient *r*_*ij*_. The functional connectivity metrics are organized into a connectivity matrix *R*, where each element *R*_*ij*_ represents the correlation *r*_*ij*_ between parcels *P*_*i*_ and *P*_*j*_.

To focus on the most significant connections, proportional thresholding is applied to the connectivity matrix. This involves retaining the top *p*% of connections, where *p* is the proportional threshold (e.g., 70%). The threshold value *τ* is determined such that only the top *p*% of the highest absolute correlation values in the connectivity matrix are retained. Let *r*_sorted_ be the sorted list of absolute correlation values from the connectivity matrix *R*. The threshold *τ* is the value at the (1− *p/*100)-th percentile of *r*_sorted_. All connections below the threshold *τ* are set to zero:

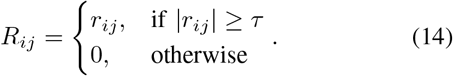

For each sequence of brain states from each functional module, *i*=1 we create a set of dynamic graphs 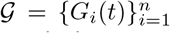, where *G*_*i*_(*t*) = (*V*_*i*_(*t*), *E*_*i*_(*t*)) represents the *i*-th dynamic graph at time snapshot *t*. For each brain state *S*_*t*_ at time *t*, the dynamic graph *G*_*i*_(*t*) is defined with vertices *V*_*i*_(*t*) representing the set of brain regions (parcels) in the *i*-th functional module, and edges *E*_*i*_(*t*) representing the set of significant functional connections between parcels, with weights corresponding to the thresholded Pearson correlation coefficients. The dynamic graph dataset is *𝒢* a sequence of graphs capturing the temporal evolution of functional connectivity within each functional module:

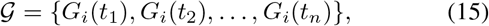

where *t*_1_, *t*_2_, …, *t*_*n*_ are the time points corresponding to different brain states.

To create a dynamic graph dataset for analysis, we compile the graphs representing each brain state into a sequence. This dynamic graph dataset is constructed to be used for subsequent classification tasks, such as disorder detection. We organize the dynamic graphs *{G*_*i*_(*t*_1_), *G*_*i*_(*t*_2_), …, *G*_*i*_(*t*_*n*_)*}* corresponding to the sequence of brain states *{t*_1_, *t*_2_, …, *t*_*n*_*}*.

### C. Dynamic Graph Transformer

#### 1) Input Embeddings

The input embeddings are crucial for capturing the complex functional connectivity patterns in the brain. Node embeddings represent the functional connectivity profile of each brain region (parcel) within the dynamic graph. These embeddings are derived from the connectivity vectors obtained from the connectivity map. For each time snapshot *t*, we construct the connectivity map *R*(*t*) for the dynamic graph *G*(*t*) = (*V* (*t*), *E*(*t*)), where *R*_*ij*_(*t*) represents the Pearson correlation coefficient between nodes *v*_*i*_ and *v*_*j*_. For each node *v*_*i*_*∈ V* (*t*), we create a connectivity vector **c**_*i*_(*t*) that captures the connectivity profile of node *v*_*i*_ with all other nodes in the graph. The connectivity vector is defined as:

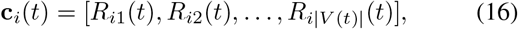

where *R*_*ij*_(*t*) is the correlation value between node *v*_*i*_ and node *v*_*j*_ at time *t*. The connectivity vector **c**_*i*_(*t*) serves as the initial node embedding for *v*_*i*_ at time *t*. These embeddings are typically passed through a learnable transformation layer, such as a linear transformation, to map them to a suitable embedding space ℝ^*d*^:

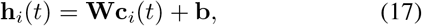

where **W***∈* ℝ^*d×*|*V* (*t*)|^ is a learnable weight matrix, **b** ∈ ℝ^*d*^ is a bias vector, and **h**_*i*_(*t*)*∈* ℝ^*d*^ is the resulting node embedding. Edge embeddings represent the strength and nature of the connections between nodes. These embeddings are based on the correlation values from the connectivity map, serving as edge features in the dynamic graph. For each edge *e*_*ij*_*∈ E*(*t*), the edge feature is derived from the correlation value *R*_*ij*_(*t*) between nodes *v*_*i*_ and *v*_*j*_. This value captures the functional connectivity strength between the nodes:

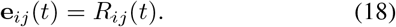

Similar to node embeddings, edge features can be transformed into a higher-dimensional embedding space through a learnable transformation. This process involves applying a nonlinear transformation to the edge feature:

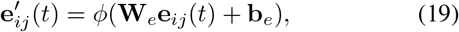

where 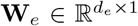 is a learnable weight matrix, 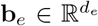 is a bias vector, *ϕ* is a non-linear activation function (e.g., ReLU), and 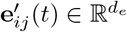 is the resulting edge embedding.

#### 2) Graph Encoder

The Graph Encoder is designed to learn and encode the structural features of dynamic brain networks. Graph Attention Networks (GATs) [44] are employed to capture the structural features of each graph snapshot *G*_*i*_(*t*) = (*V*_*i*_(*t*), *E*_*i*_(*t*)). GATs utilize attention mechanisms to weigh the importance of neighboring nodes and edges when updating node embeddings, allowing for more expressive representations of graph structure. Let **h**_*i*_(*t*) *∈*ℝ^*d*^ represent the embedding of node *v*_*i*_ at time *t*. Initially, these embeddings are derived from the connectivity vectors as described in the input embeddings section. For each node *v*_*i*_, the attention mechanism computes attention coefficients *α*_*ij*_ for its neighboring nodes *v*_*j*_*∈ N* (*v*_*i*_), where *N*(*v*_*i*_) denotes the set of neighbors of *v*_*i*_. The attention scores *e*_*ij*_ are calculated as:

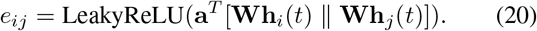

The attention coefficients *α*_*ij*_ are then obtained by normalizing these scores using the softmax function:

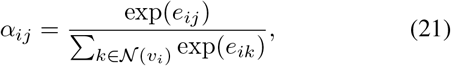

where **W** is a weight matrix, ∥ denotes concatenation, **a** is a learnable attention vector, and LeakyReLU is the leaky rectified linear unit activation function. The node embeddings are then updated by aggregating the embeddings of neighboring nodes, weighted by the attention coefficients:

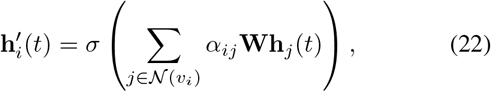

where *σ* is an activation function, such as ReLU. To stabilize the learning process and improve representation power, multihead attention is applied. This involves executing the attention mechanism *K* times with different learnable parameters and concatenating the results:

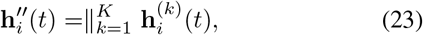

where 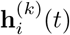 is the updated embedding from the *k*-th attention head.

To obtain fixed-size representations of each graph snapshot *G*_*i*_(*t*), graph pooling (mean pooling) is applied to the node embeddings produced by the GAT. Mean pooling computes the average of all node embeddings in the graph snapshot, resulting in a fixed-size representation irrespective of the graph size:

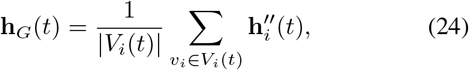

where **h**_*G*_(*t*) *∈* ℝ^*Kd*^ is the pooled graph embedding, |*V*_*i*_(*t*)| is the number of nodes in the graph snapshot, and *Kd* is the dimension of the concatenated embeddings from multi-head attention. The result of mean pooling is a fixed-size vector that represents the entire graph snapshot, capturing the aggregate structural information learned by the GAT.

#### 3) Temporal Encoder

The Temporal Encoder is designed to capture the temporal dynamics of brain connectivity by processing sequences of graph snapshots. The Temporal Encoder processes sequences of graph snapshots *{G*_*i*_(*t*_1_), *G*_*i*_(*t*_2_), …, *G*_*i*_(*t*_*n*_) to capture temporal dependencies. These sequences may vary in length, necessitating padding and masking to ensure uniform input sizes. Shorter sequences are padded with zero vectors to match the length of the longest sequence in the batch. A binary mask differentiates between actual data and padding:

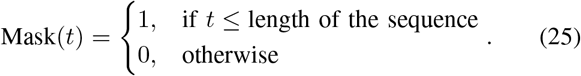

To incorporate the temporal order of graph snapshots, positional encoding is added to the graph embeddings. Positional encoding provides a way to inject information about the position of each snapshot in the sequence.

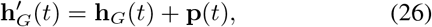

where 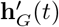 is the positionally encoded graph embedding. The multi-head attention mechanism is used to model the dependencies between graph snapshots at different time points [45]. For each pair of snapshots *G*_*i*_(*t*) and *G*_*i*_(*t*^*′*^), the attention score is calculated using the scaled dot-product attention:

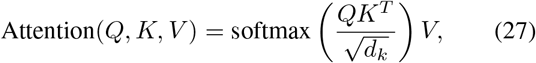

where *Q, K*, and *V* are the query, key, and value matrices derived from the input embeddings. Multiple attention heads are used to jointly attend to information from different representation subspaces. The output of the multi-head attention is passed through position-wise feed-forward layers to introduce non-linearity and further transform the embeddings. Each feedforward layer consists of two linear transformations with a ReLU activation in between:

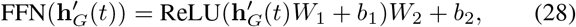

where *W*_1_, *W*_2_, *b*_1_, and *b*_2_ are learnable parameters. The output is normalized to stabilize the training process. To obtain a fixed-size representation of the entire dynamic graph, temporal pooling is applied to the transformed graph embeddings. Mean pooling aggregates the embeddings across all time points to produce a single representation:

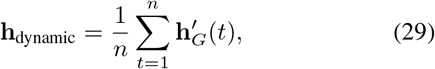

where *n* is the number of graph snapshots in the sequence. The result of mean pooling is a fixed-size vector that represents the entire sequence of graph snapshots, capturing the temporal dynamics of brain connectivity.

#### 4) Adaptive Fusion

The Adaptive Fusion process in the BrainDGT is critical for integrating the spatio-temporal features captured by multiple temporal encoders into a single, cohesive representation. The Spatio-temporal features are extracted through a series of temporal encoders applied to dynamic graph sequences. Each temporal encoder processes sequences of graph snapshots, capturing the evolution of connectivity patterns over time. Let 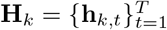 represent the set of spatio-temporal features extracted by the *k*-th temporal encoder, where **h**_*k,t*_*∈* ℝ^*d*^ is the feature vector for the *k*-th encoder at time *t*, and *T* is the number of time steps. The core of the adaptive fusion process is an attention mechanism designed to weigh the spatio-temporal features based on their relevance. This mechanism ensures that the final integrated representation is both comprehensive and focused on the most informative aspects of the data. The spatio-temporal features from all temporal encoders are concatenated to form a comprehensive feature matrix. Let *K* be the number of temporal encoders:

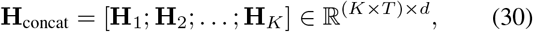

where [.; .] denotes concatenation along the temporal dimension. Construct the query *Q*, key *K*, and value *V* matrices for the attention mechanism from the concatenated feature matrix **H**_concat_:

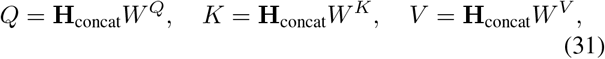

where *W*^*Q*^, *W*^*K*^, *W*^*V*^ *∈* ℝ^*d×d*^ are learnable weight matrices. Compute the attention scores using the scaled dot-product attention mechanism. Apply the attention scores to the value matrix *V* to obtain the weighted feature matrix:

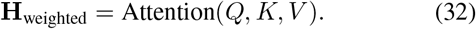

The weighted feature matrix **H**_weighted_ is then aggregated to form a single representation that captures the essential spatio-temporal dynamics of the brain network. Perform global pooling (e.g., mean pooling) across the temporal dimension to integrate the weighted features into a fixed-size representation:

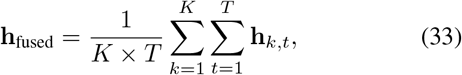

where **h**_fused_ *∈*ℝ^*d*^ is the final integrated representation.

#### 5) Classification

The classification process use the fused spatio-temporal features to perform disorder classification. The fused features obtained from the Adaptive Fusion process represent a comprehensive summary of the dynamic spatiotemporal brain connectivity. These features serve as input to a classification network to predict disorder states. Let **h**_fused_ *∈* ℝ^*d*^ be the fused feature vector, which captures the essential information needed for classification. The classification network consists of fully connected layers organized in a Multilayer Perceptron (MLP). These layers transform the fused feature vector into class probabilities. The MLP comprises several fully connected layers, each followed by a non-linear activation function such as ReLU. Let **h**_0_ = **h**_fused_. The transformation through the *l*-th layer is given by:

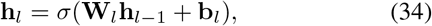

where 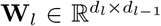 is the weight matrix, 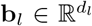 is the bias vector, *σ* is an activation function (e.g., ReLU), and *d*_*l*_ is the number of neurons in the *l*-th layer. The final fully connected layer maps the transformed features to class scores. For binary classification, this layer has one output neuron. For binary classification, a sigmoid activation function is applied to the output neuron to produce a probability score for the positive class. The probability of the positive class for binary classification is given by:

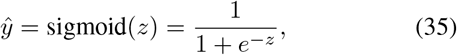

where *z* is the output score from the final layer. The crossentropy loss function measures the discrepancy between the predicted probabilities and the true labels. For binary classification, the loss is defined as:

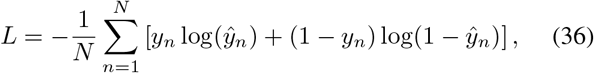

where *N* is the number of samples, *y*_*n*_ is the true label, and *ŷ*_*n*_ is the predicted probability for sample *n*. The Adam optimizer is used to minimize the cross-entropy loss and update the model parameters. The parameter updates are computed as:

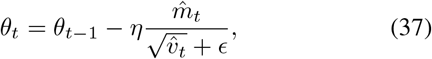

where *θ*_*t*_ represents the model parameters at iteration *t, η* is the learning rate, 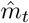 and 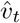 are bias-corrected first and second moment estimates, respectively, and *ϵ* is a small constant for numerical stability. Learning rate scheduling adjusts the learning rate during training to improve convergence. The pseudocode of BrainDGT is shown in Algorithm 1.

## IV. Experimental Results

In this section, we evaluate the effectiveness of our proposed BrainDGT model through comprehensive experiments. Our evaluation focuses on addressing the following research questions:

RQ1: How accurately does BrainDGT diagnose brain disorders using real fMRI data, and what classification improvements does BrainDGT offer over state-of-the-art sliding window methods and other baselines?

RQ2: What is the impact of removing specific components of the BrainDGT model on its overall performance?

RQ3: What are the optimal parameters for the BrainDGT model to achieve the highest diagnostic accuracy?

### A. Experimental Settings

#### 1) Datasets

##### a) ADNI Dataset

We used the Alzheimer’s Disease Neuroimaging Initiative (ADNI) dataset, available through the ADNI database^2^. This dataset provides fMRI data crucial for studying Alzheimer’s Disease (AD), focusing on identifying biomarkers for early detection and monitoring. Subjects are categorized as either cognitively normal or diagnosed with AD. From this dataset, we select a subset of 426 subjects with resting-state fMRI data, comprising 199 women (46.7%) and 146 individuals with AD (34.2%), aged 50 to 100 years. Rigorous selection criteria ensure accurate diagnoses, enhancing the dataset’s reliability.

###### Algorithm 1

BrainDGT

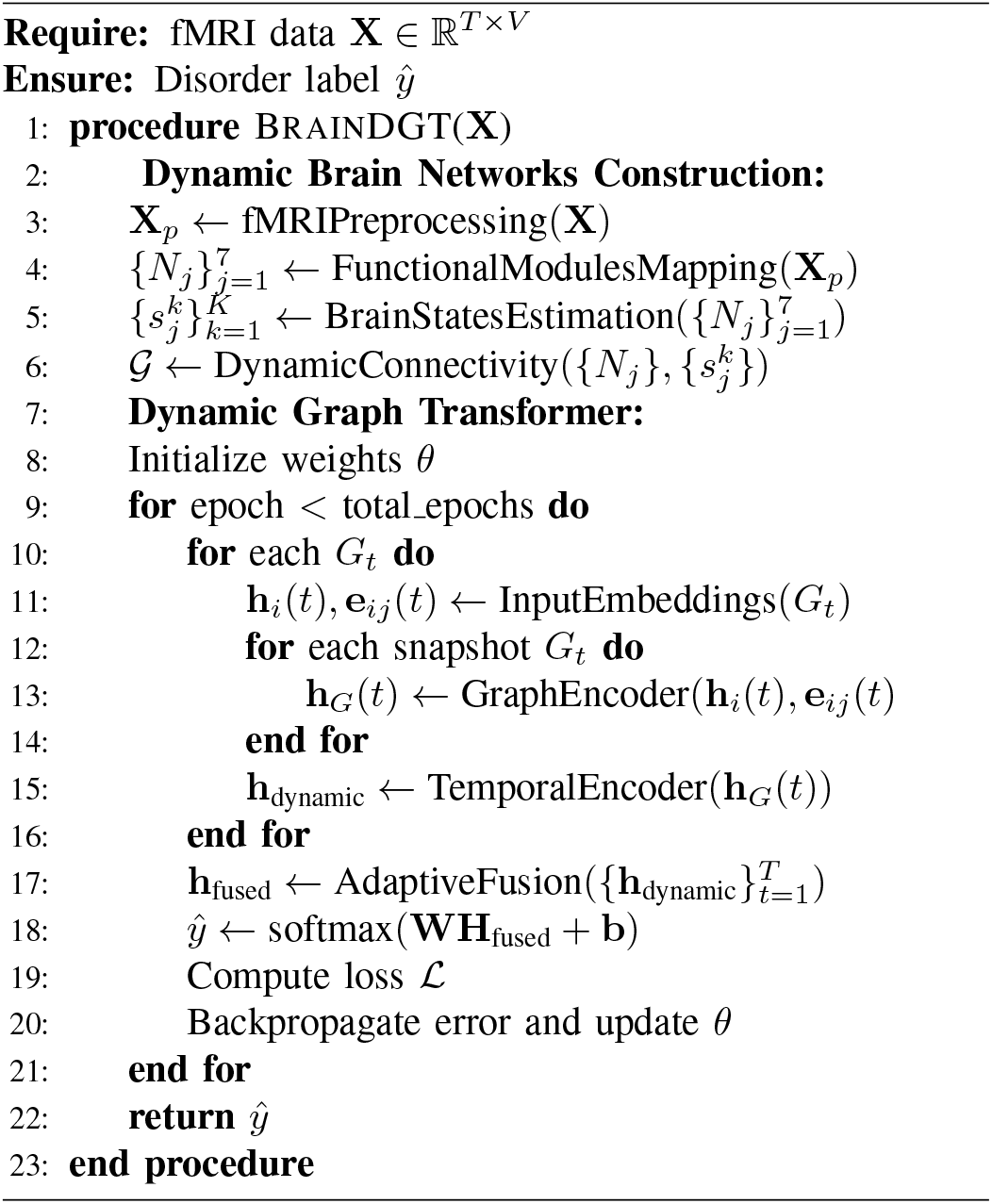

##### b) PPMI Dataset

The Parkinson’s Progression Markers Initiative (PPMI) dataset, accessible via the PPMI website^3^, includes extensive fMRI data for tracking Parkinson’s Disease (PD) progression. Researchers access this data after meeting specific criteria, facilitating in-depth studies on PD biomarkers. The dataset’s primary objective is to identify diagnostic and progression markers for PD. We select a cohort of 823 subjects, including 230 PD patients (70.9%) and 130 females (40.1%), aged 40 to 85 years. This representative sample enhances the dataset’s utility for PD research.

##### c) ABIDE Dataset

The Autism Brain Imaging Data Exchange (ABIDE) dataset, provided by the ABIDE consortium^4^, offers a comprehensive collection of fMRI data to study autism spectrum disorder (ASD). This publicly accessible dataset includes contributions from various international sites, aimed at exploring the neural basis of ASD and developing diagnostic markers. For our analysis, we select a subset of 1118 subjects with fMRI data from social cognition tasks, including 537 individuals diagnosed with ASD (48%) and 161 females (14.5%), aged 8 to 40 years. This diverse dataset supports extensive research on neural characteristics and potential diagnostic markers of ASD. A detailed summary of the datasets is presented in Table I.

**TABLE I.** Statistices of Datasets.

| Dataset | Subjects | Age Range | Male/Female | Labels |
| --- | --- | --- | --- | --- |
| ADNI | 427 | 50-100 | 227/199 | CN (280), AD (146) |
| PPMI | 324 | 40-85 | 194/130 | Control (94), PD (230) |
| ABIDE | 1118 | 8-40 | 957/161 | Control (581), ASD (537) |

#### 2) Baselines

We selected a diverse range of state-of-the-art methods as baselines to ensure a comprehensive comparison with BrainDGT. These baselines encompass traditional machine learning and advanced dynamic brain network models, providing a holistic evaluation of BrainDGT’s performance.

##### a) Conventional Machine Learning Models

We include Support Vector Machine (SVM) [46], Random Forest [47], and Multi-Layer Perceptron (MLP) [48] as representatives of conventional machine learning techniques. These models are trained on features extracted from brain networks constructed using the BrainGB pipeline. The features, derived through graph theory methods, encapsulate the structural properties of the brain networks. The scikit-learn library is used to implement these models. Their selection is based on their extensive application and proven effectiveness in various classification tasks, including neuroimaging data.

##### b) Graph Learning Models

We include Graph Convolutional Network (GCN) [49], Graph Isomorphism Network (GIN) [50], and Graph Attention Network (GAT) [44] to represent graph learning approaches. Brain networks are constructed from fMRI data using the BrainGB pipeline and directly applied to these graph learning models without feature selection. The PyTorch Geometric library is used for implementation. These models are chosen for their ability to capture complex relationships and interactions within graph-structured data, which is crucial for accurate brain activity analysis.

##### c) Static Brain Network Models

We include BrainGNN [23], BrainGB [4], and BRAINNETTF [24] as static brain network models. These models are trained according to their respective implementations, focusing on capturing static connectivity patterns within the brain. Their selection is based on their specific design for handling brain network data and their previous success in brain disorder diagnosis tasks.

##### d) Basic Dynamic Brain Network Models

We include STfMRI [30], ST-GCNs [29], and STAGIN [15] as basic dynamic brain network models. These models utilize fixed sliding windows to extract dynamic functional connectivity from fMRI data, generally focusing on phenotype prediction and leveraging graph convolutional networks as their core technology. They provide a baseline for dynamic modeling capabilities using conventional techniques.

##### e) Advanced Dynamic Brain Network Models

We include DART [16], MDGL [37], and OT-MCSTGCN [38] as advanced dynamic brain network models. These models also employ fixed sliding windows but incorporate advanced techniques such as Transformers and attention mechanisms to capture dynamic and heterogeneous brain activity. Specifically designed for brain disorder diagnosis, these models represent the state of the art in dynamic brain network modeling.

#### 3) Implementation Details

We implemented BrainDGT in Python with PyTorch to capture dynamic brain activity from fMRI data. The preprocessing refines raw fMRI data into functional ROIs using the Schaefer Atlas, detects neural events via HRF estimation, and constructs dynamic brain networks as spatio-temporal graphs. The Dynamic Graph Transformer employs Graph Attention Networks (GATs) for spatial dependencies and Transformer encoders for temporal dependencies, followed by adaptive fusion and classification with a fully connected neural network. The model is trained and evaluated on three fMRI datasets, partitioned into 70% training, 15% validation, and 15% testing subsets. Training uses the Adam optimizer with a 0.001 learning rate, crossentropy loss, and early stopping based on validation loss. Hyperparameters are tuned through grid search and crossvalidation, identifying optimal values of three GAT layers and two Transformer layers. The window size for segmenting neural event sequences uses HRF estimation. Computational resources include an NVIDIA GeForce RTX 4090 GPU, 64GB of RAM, and an Intel Core i9 processor, ensuring efficient handling of BrainDGT computations.

#### 4) Evaluation Metrics

We evaluated BrainDGT and the baseline methods using three key metrics: accuracy, F1-score, and the area under the ROC curve (AUC).

#### 5) Computational Complexity

We assessed the efficiency of the BrainDGT model by analyzing its computational complexity. The time complexity is mainly due to the graph encoder and the temporal encoder. The graph encoder, using graph attention networks (GAT), has a time complexity of *O*(*N*^2^), where *N* is the number of nodes. The temporal encoder, using Transformer encoders, has a complexity of *O*(*n*^2^ *·d*), where *n* is the sequence length and *d* is the dimension of the model. The space complexity considers the storage of dynamic brain networks and model parameters: *O*(*N*^2^ *·T*) for the networks, with *N* nodes and *T* time points, and *O*(*D*) for the parameters, where *D* is the total number of parameters. Despite its high computational complexity, BrainDGT effectively captures dynamic brain activity and is efficient in detecting brain disorders, justifying its computational cost.

### B. Performance of BrainDGT

We evaluated BrainDGT using the ADNI, PPMI, and ABIDE datasets, which represent various subject demographics and brain disorder classifications, providing a comprehensive assessment of the model’s generalizability. BrainDGT achieves the highest accuracy on the PPMI dataset (88.58%±4.43), indicating effective capture of Parkinson’s Disease (PD) neural patterns, although the F1 score is lower (62.63%±1.54) due to variability in PD symptoms. On the ABIDE dataset, BrainDGT shows a high AUC (88.30%±3.97), reflecting excellent discriminative power, but has slightly lower accuracy and F1 scores, highlighting challenges in distinguishing Autism Spectrum Disorder (ASD) from control subjects.

The ADNI dataset shows moderate performance with an accuracy of 77.52%±1.89, F1 score of 72.57%±4.81, and AUC of 80.02%±1.98, indicating that while BrainDGT can identify Alzheimer’s Disease (AD) neural activity, improvement is needed in reducing false positives and enhancing sensitivity.

### C. Comparison with Baseline Methods (RQ1)

We compare BrainDGT with various baseline methods using the ADNI, PPMI, and ABIDE datasets. Baseline methods are grouped into categories. For each group, we analyze BrainDGT’s performance relative to these baselines and discuss the reasons for observed differences. Table II summarizes the results, highlighting BrainDGT’s superior performance.

**TABLE II.** Comparative Performance Analysis (%). BrainDGT demonstrates statistically significant improvements over baseline models confirmed by t-tests (p-value ***<*** 0.05).

| Baseline Group | Method | ADNI |  |  | PPMI |  |  | ABIDE |  |  |
| --- | --- | --- | --- | --- | --- | --- | --- | --- | --- | --- |
|  |  | Accuracy | F1 | AUC | Accuracy | F1 | AUC | Accuracy | F1 | AUC |
| Machine Learning | SVM | 60.47±3.34 | 43.79±0.67 | 64.57±0.97 | 63.65±1.31 | 36.62±4.57 | 41.61±2.74 | 63.90±4.98 | 40.60±1.50 | 65.62±0.90 |
|  | Random Forest | 60.65±4.31 | 44.47±4.96 | 63.98±2.36 | 65.36±1.00 | 37.75±3.21 | 41.22±3.66 | 64.80±0.67 | 41.75±2.27 | 67.81±4.69 |
|  | MLP | 61.04±4.75 | 45.46±1.99 | 66.22±2.79 | 66.14±3.93 | 36.48±1.63 | 42.19±3.71 | 64.91±0.86 | 42.65±0.58 | 68.00±2.11 |
| Graph Learning | GCN | 65.74±4.52 | 52.81±1.29 | 69.92±4.75 | 72.81±0.96 | 40.86±4.36 | 45.76±1.93 | 67.85±3.18 | 50.94±2.09 | 71.86±1.54 |
|  | GIN | 64.86±3.05 | 54.90±4.29 | 68.49±2.04 | 75.06±3.94 | 41.85±2.47 | 46.85±3.28 | 69.14±3.75 | 51.68±2.81 | 72.66±0.93 |
|  | GAT | 66.23±3.04 | 55.52±4.85 | 69.25±1.06 | 74.85±4.10 | 42.38±4.95 | 47.41±1.68 | 71.18±3.52 | 53.98±3.20 | 74.98±4.45 |
| Static Brain Network Models | BrainGNN | 68.57±1.14 | 56.72±1.32 | 72.88±0.88 | 77.57±4.10 | 45.58±2.66 | 49.87±1.44 | 73.70±4.15 | 55.32±2.27 | 75.16±4.48 |
|  | BrainGB | 69.82±1.81 | 57.06±2.10 | 73.99±1.40 | 79.24±1.51 | 46.42±0.53 | 51.31±4.82 | 74.18±3.22 | 56.94±3.20 | 76.70±1.86 |
|  | BRAINNETTF | 70.82±2.60 | 57.82±0.06 | 74.91±4.96 | 80.21±2.17 | 47.07±0.90 | 52.18±4.53 | 75.25±1.01 | 57.86±4.62 | 77.38±1.19 |
| Dynamic brain network models (Basic) | ST-fMRI | 69.76±4.67 | 58.57±3.50 | 70.18±1.16 | 81.99±4.19 | 49.76±3.06 | 54.48±5.00 | 76.57±2.82 | 59.18±0.73 | 81.89±2.81 |
|  | ST-GCNs | 71.49±3.81 | 61.07±3.39 | 72.81±3.68 | 82.56±4.81 | 51.26±4.35 | 56.02±4.20 | 78.52±3.45 | 61.01±4.00 | 82.50±1.96 |
|  | STAGIN | 72.25±3.83 | 63.30±2.08 | 73.73±3.46 | 83.17±2.68 | 52.21±2.13 | 57.97±1.72 | 80.16±1.83 | 63.38±2.36 | 83.24±1.38 |
| Dynamic brain network models (Advanced) | DART | 72.76± 1.2 | 62.31±1.22 | 70.91±0.63 | 81.77±3.47 | 55.34±4.70 | 64.22±1.55 | 82.53±2.26 | 64.24±3.80 | 84.17±2.60 |
|  | MDGL | 73.59±1.91 | 67.40±4.50 | 74.76±1.05 | 83.89±4.59 | 59.17±4.19 | 65.51±3.34 | 82.86±3.28 | 65.45±3.20 | 85.25±4.68 |
|  | OT-MCSTGCN | 74.92±3.03 | 69.36±1.15 | 75.29±1.00 | 84.85±3.48 | 60.87±3.77 | 66.41±3.98 | 83.60±0.58 | 65.98±4.28 | 86.43±2.55 |
| Ours | BrainDGT | **77.52±1.89** | **72.57±4.81** | **80.02±1.98** | **88.58±4.43** | **62.63±1.54** | **68.38±3.92** | **85.42±2.51** | **68.59±3.10** | **88.30±3.97** |

*Conventional machine learning models* used for comparison include SVM, RF, and MLP. These models employ graph theory-based features extracted from brain networks constructed using the BrainGB pipeline. Our BrainDGT model significantly outperforms the best performing conventional machine learning model, MLP, across all datasets. In the ADNI dataset, BrainDGT’s accuracy of 77.52% is 27.0% higher than MLP’s 61.04%. The F1 score for BrainDGT is 59.5% higher than MLP’s 45.46%, and AUC is 20.9% higher than MLP’s 66.22%. Similar improvements are observed in the PPMI and ABIDE datasets, with BrainDGT consistently achieving higher scores in all metrics. The superior performance of BrainDGT can be attributed to its ability to capture the dynamic and heterogeneous nature of brain activity, which conventional machine learning models fail to do. By using adaptive segmentation of ROIs and dynamic graph representations, BrainDGT provides a more accurate and nuanced understanding of brain connectivity patterns. This approach overcomes the limitations of fixed sliding windows and static feature extraction methods, leading to more reliable and precise predictions.

*Graph learning models*, like GCN, GIN, and GAT, apply graph techniques to fMRI brain networks. BrainDGT outperforms GAT. On ADNI, BrainDGT’s accuracy is 17.0% higher than GAT’s 66.23%, its F1 score is 30.6% higher than GAT’s 55.52%, and its AUC is 15.6% higher than GAT’s 69.25%. Similar improvements are seen on PPMI and ABIDE datasets. This superior performance is due to BrainDGT’s dynamic approach, capturing temporal variations in brain connectivity, unlike static models like GCN, GIN, and GAT. BrainDGT’s temporal encoders and adaptive fusion mechanisms enable it to better track brain activity evolution, resulting in more accurate predictions.

*Static brain network models* in our comparison include BrainGNN, BrainGB, and BRAINNETTF. These static models extract features that do not change over time. Our BrainDGT model consistently outperforms BRAINNETTF across all datasets. In ADNI, BrainDGT’s accuracy is 9.4% higher than BRAINNETTF’s 70.82%, F1 score is 25.5% higher than BRAINNETTF’s 57.82%, and AUC is 6.8% higher than BRAINNETTF’s 74.91%. Similar results are in PPMI and ABIDE. Static models fail to capture fMRI temporal dynamics. BrainDGT models both spatial and temporal variations, providing a more comprehensive analysis. Dynamic graphs allow BrainDGT to identify transient changes in connectivity, leading to more accurate and reliable disorder detection.

*Basic dynamic brain network models* like ST-fMRI, STGCNs, and STAGIN use fixed sliding windows for dynamic functional connectivity from fMRI data and graph learning techniques. BrainDGT significantly outperforms STAGIN. In the ADNI dataset, BrainDGT’s accuracy is 5.2% higher, F1 score is 14.6% higher, and AUC is 8.6% higher than STAGIN. BrainDGT also consistently outperforms ST-fMRI, ST-GCN, and STAGIN in PPMI and ABIDE datasets. Fixed sliding windows in basic models may capture brain activity dynamics suboptimally. BrainDGT overcomes this by adaptively segmenting BOLD signals based on HRF estimation, accurately representing dynamic brain connectivity. This, along with advanced graph and temporal encoders, enables superior performance.

*Advanced dynamic brain network models*, such as DART, MDGL, and OT-MCSTGCN, use techniques like transformers and attention mechanisms. BrainDGT surpasses OTMCSTGCN in all metrics: 3.5% higher accuracy (74.92% vs. OT-MCSTGCN), 4.6% higher F1 score (69.36%), and 4.7% higher AUC (75.29%) on the ADNI dataset. BrainDGT shows similar superior performance on PPMI and ABIDE datasets. While OT-MCSTGCN relies on fixed sliding windows, BrainDGT’s adaptive segmentation and fusion of dynamic/static features offer a more accurate view of brain activity. Its advanced graph-temporal encoders capture complex brain dependencies and interactions, improving disorder detection.

### D. Ablation Study (RQ2)

We conducted an ablation study on BrainDGT to evaluate the contribution of its components. The baseline model includes all components from the methods section. We created several scenarios by removing or altering components. First, we evaluated the impact of using raw fMRI data by omitting preprocessing. Then, we assessed heterogeneity by constructing 400-node brain networks without further segmentation. To analyze dynamic modeling, we replaced HRF-based sequences with fixed sliding windows. We also omitted the graph and temporal encoders to understand their impact. Finally, we removed the adaptive fusion process to evaluate its role. We used consistent datasets (ADNI, PPMI, ABIDE) and data splits for training, validation, and testing. Multiple iterations determined the statistical significance of differences. Each model’s performance is compared to the baseline using accuracy, F1-score, and AUC, and results are in Fig. 4.

**Fig. 4.**
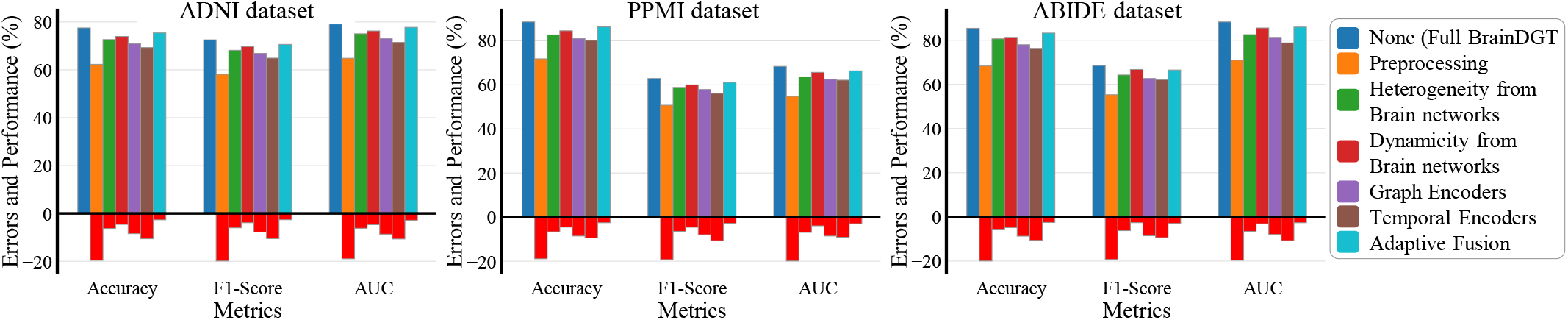
Ablation studies of BrainDGT.

The results of the ablation study show a significant performance drop when key components are removed from BrainDGT. The removal of pre-processing causes the largest drop, highlighting its role in enhancing signal quality and reducing noise. The Accuracy on the ADNI dataset drops by 19.2%, with F1 score and AUC dropping by 19.4% and 15.17%, respectively. Omitting network heterogeneity leads to notable declines, with accuracy drops of 4.9% (ADNI) and 4.8% (ABIDE), underscoring the importance of modeling heterogeneous activation patterns. Removing dynamic modeling decreases accuracy by 3.5% to 4.1%, showing the need to reflect fluctuations in temporal neural activity. Excluding graph and temporal encoders, crucial for capturing structural and temporal features, results in 6.3% to 7.2% and 8.2% to 8.4% accuracy drops, respectively. Lastly, removing adaptive fusion, which integrates these features for robust detection of brain disorders, causes marked performance degradations.

### E. Parameter Analysis (RQ3)

We performed parameter analysis to determine the impact of key variables on BrainDGT model performance in detecting brain disorders using fMRI data. Specifically, we analyzed the number of functional ROIs (*n*) and the number of layers (*l*) in the graph and temporal encoders. These parameters were varied to find their optimal values for model accuracy and reliability. The number of functional ROIs (*n*) ranged from 1 to 17 using predefined networks within Schaefer atlases. The number of layers (*l*) varied from 1 to 5 to capture the complexity and temporal dependencies of neural activity.

We performed experiments using a grid where each point represents a unique combination of *n* and *l*. These experiments are performed on the ADNI, PPMI, and ABIDE fMRI datasets, keeping all other variables constant. BrainDGT is trained and evaluated using each parameter combination. Our analysis reveals distinct patterns for each dataset. For the ADNI dataset, the highest precision (77.52%) and F1 score (72.57%) are achieved with *n* = 10 and *l* = 2. Increasing ROIs beyond 10 decreases performance, suggesting an optimal granularity level. A shallow network with 2 layers yields the best results, with deeper networks not significantly improving performance. In the PPMI dataset, optimal performance is achieved with *n* = 9 and *l* = 2, resulting in 88.58% accuracy and a 62.63% F1 score. Similar to ADNI, more than 9 ROIs lead to diminishing returns, and the 2-layer depth provides the best balance between complexity and performance. For the ABIDE dataset, the best results are obtained with *n* = 7 and *l* = 3, achieving 85.42% accuracy and a 68.59% F1 score, indicating a slightly deeper network is beneficial. Figs. 5 and 6 depict these relationships, showing trends in accuracy and F1 score with varying *n* and *l*.

**Fig. 5.**
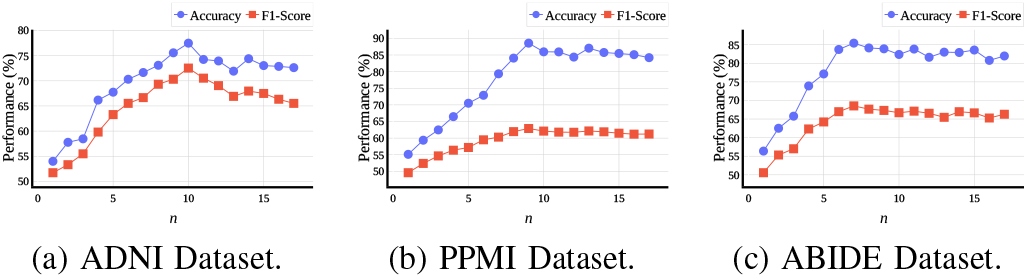
The accuracy and F1-score of BrainDGT w.r.t. different ***n*** values.

**Fig. 6.**
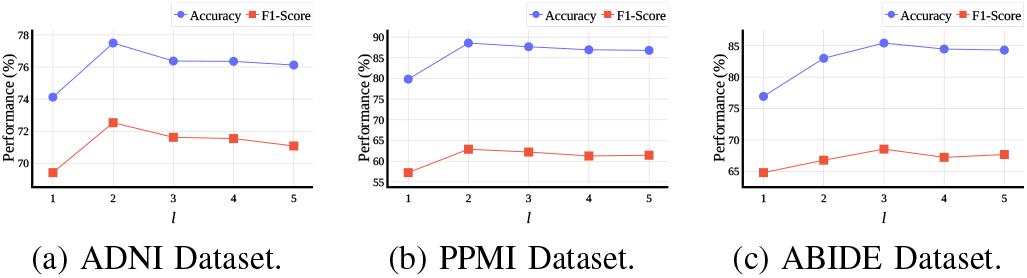
The accuracy and F1-score of BrainDGT w.r.t. different ***l*** values.

## V. CONCLUSION

In this article, we study dynamic brain networks, which play a major part in diagnosing brain disorders. We propose BrainDGT, a dynamic Graph Transformer model for analyzing modular brain activities. The BrainDGT model improves the accuracy of brain disorder diagnosis from three perspectives, including adaptive temporal length estimation, modular segmentation of fMRI scans, and dual attention mechanisms. First, compared with other models that utilize fixed temporal lengths, BrainDGT estimates adaptive brain states through deconvolution of the Hemodynamic Response Function (HRF), avoiding the constraints of fixed-size windows. Second, BrainDGT effectively segments fMRI scans into functional modules based on established brain networks, allowing for detailed, module-specific analysis. Additionally, the BrainDGT model introduces a dual attention mechanism: graph-attention extracts structural features from dynamic brain network snapshots, while self-attention identifies significant temporal dependencies. These spatio-temporal features are adaptively fused into a unified representation for disorder classification.

Experimental results show that the proposed model outperforms the baselines on three real fMRI datasets, ADNI, PPMI, and ABIDE, proving that BrainDGT can effectively classify brain network representations. In summary, our study realizes the improved diagnosis of brain disorders and promotes the use of dynamic Graph Transformer technology in brain analysis. In the future, we will refine the model and explore its application to other neuroimaging data to enhance its clinical utility and effectiveness.

## Data Availability

All data produced in the present study are available upon reasonable request to the authors

## Footnotes

1 https://github.com/ThomasYeoLab/CBIG/tree/master/stable_projects/brain_parcellation/Schaefer2018_LocalGlobal

2 http://adni.loni.usc.edu/data-samples/access-data

3 https://www.ppmi-info.org/access-data-specimens/download-data

4 http://fcon_1000.projects.nitrc.org/indi/abide

## References

[1] M. Hallett, W. De Haan, G. Deco, R. Dengler, R. Di Iorio, C. Gallea, C. Gerloff, C. Grefkes, R. C. Helmich, M. L. Kringelbach, F. Miraglia, I. Rektor, O. Strýček, F. Vecchio, L. J. Volz, T. Wu, and P. M. Rossini, “Human brain connectivity: Clinical applications for clinical neurophysiology,” Clinical Neurophysiology, vol. 131, no. 7, pp. 1621–1651, Jul. 2020.

[2] Q. Yu, Y. Du, J. Chen, J. Sui, T. Adali, G. D. Pearlson, and V. D. Calhoun, “Application of Graph Theory to Assess Static and Dynamic Brain Connectivity: Approaches for Building Brain Graphs,” Proceedings of the IEEE, vol. 106, no. 5, pp. 886–906, May 2018.

[3] M. R. Ahmed, Y. Zhang, Z. Feng, B. Lo, O. T. Inan, and H. Liao, “Neuroimaging and machine learning for dementia diagnosis: Recent advancements and future prospects,” IEEE Reviews in Biomedical Engineering, vol. 12, pp. 19–33, 2019.

[4] H. Cui, W. Dai, Y. Zhu, X. Kan, A. A. C. Gu, J. Lukemire, L. Zhan, L. He, Y. Guo, and C. Yang, “BrainGB: A Benchmark for Brain Network Analysis With Graph Neural Networks,” IEEE Transactions on Medical Imaging, vol. 42, no. 2, pp. 493–506, Feb. 2023.

[5] C. Peng, M. Liu, C. Meng, S. Yu, and F. Xia, “Adaptive brain network augmentation based on group-aware graph learning,” in International Conference on Learning Representations (ICLR), Vienna, Austria, May 2024.

[6] Z. Wang, J. Xin, Z. Wang, Y. Yao, Y. Zhao, and W. Qian, “Brain functional network modeling and analysis based on fMRI: A systematic review,” Cognitive Neurodynamics, vol. 15, no. 3, pp. 389–403, Jun. 2021.

[7] E. Bullmore and O. Sporns, “Complex brain networks: graph theoretical analysis of structural and functional systems,” Nat. Rev. Neurosci., vol. 10, no. 3, pp. 186–198, Feb. 2009.

[8] A. Bessadok, M. A. Mahjoub, and I. Rekik, “Graph Neural Networks in Network Neuroscience,” IEEE Transactions on Pattern Analysis and Machine Intelligence, vol. 45, no. 5, pp. 5833–5848, May 2023.

[9] F. I. Karahanoğlu and D. Van De Ville, “Dynamics of large-scale fmri networks: Deconstruct brain activity to build better models of brain function,” Current Opinion in Biomedical Engineering, vol. 3, pp. 28–36, Sep. 2017.

[10] J. Skarding, B. Gabrys, and K. Musial, “Foundations and modeling of dynamic networks using dynamic graph neural networks: A survey,” IEEE Access, vol. 9, pp. 79 143–79 168, 2021.

[11] B. Hu, Z.-H. Guan, G. Chen, and C. L. P. Chen, “Neuroscience and network dynamics toward brain-inspired intelligence,” IEEE Transactions on Cybernetics, vol. 52, no. 10, pp. 10 214–10 227, Oct. 2022.

[12] A. Shehzad, F. Xia, S. Abid, C. Peng, S. Yu, D. Zhang, and K. Verspoor, “Graph transformers: A survey,” 2024.

[13] C. Ying, T. Cai, S. Luo, S. Zheng, G. Ke, D. He, Y. Shen, and T.-Y. Liu, “Do transformers really perform badly for graph representation?” in Advances in Neural Information Processing Systems, M. Ranzato, A. Beygelzimer, Y. Dauphin, P. Liang, and J. W. Vaughan, Eds., vol. 34. Curran Associates, Inc., 2021, pp. 28 877–28 888.

[14] D. Kreuzer, D. Beaini, W. Hamilton, V. Létourneau, and P. Tossou, “Rethinking graph transformers with spectral attention,” in Advances in Neural Information Processing Systems, M. Ranzato, A. Beygelzimer, Y. Dauphin, P. Liang, and J. W. Vaughan, Eds., vol. 34. Curran Associates, Inc., 2021, pp. 21 618–21 629.

[15] B.-H. Kim, J. C. Ye, and J.-J. Kim, “Learning dynamic graph representation of brain connectome with spatio-temporal attention,” in Annual Conference on Neural Information Processing Systems 2021, NeurIPS, Virtual, Dec. 2021, pp. 4314–4327.

[16] X. Kan, A. A. C. Gu, H. Cui, Y. Guo, and C. Yang, “Dynamic brain transformer with multi-level attention for functional brain network analysis,” in IEEE EMBS International Conference on Biomedical and Health Informatics (BHI). Pittsburgh, PA, USA: IEEE, Oct. 2023, pp. 1–4.

[17] C. Seguin, O. Sporns, and A. Zalesky, “Brain network communication: Concepts, models and applications,” Nature Reviews Neuroscience, vol. 24, no. 9, pp. 557–574, Sep. 2023.

[18] Y. Du, S. Fang, X. He, and V. D. Calhoun, “A survey of brain functional network extraction methods using fmri data,” Trends in Neurosciences, p. S0166223624000912, Jun. 2024.

[19] D. Sadeghi, A. Shoeibi, N. Ghassemi, P. Moridian, A. Khadem, R. Alizadehsani, M. Teshnehlab, J. M. Gorriz, F. Khozeimeh, Y.-D. Zhang, S. Nahavandi, and U. R. Acharya, “An overview of artificial intelligence techniques for diagnosis of Schizophrenia based on magnetic resonance imaging modalities: Methods, challenges, and future works,” Computers in Biology and Medicine, vol. 146, p. 105554, Jul. 2022.

[20] J. Kawahara, C. J. Brown, S. P. Miller, B. G. Booth, V. Chau, R. E. Grunau, J. G. Zwicker, and G. Hamarneh, “BrainNetCNN: Convolutional neural networks for brain networks; towards predicting neurodevelopment,” NeuroImage, vol. 146, pp. 1038–1049, Feb. 2017.

[21] A. D. Arya, S. S. Verma, P. Chakarabarti, T. Chakrabarti, A. A. Elngar, A.-M. Kamali, and M. Nami, “A systematic review on machine learning and deep learning techniques in the effective diagnosis of Alzheimer’s disease,” Brain Informatics, vol. 10, no. 1, p. 17, Dec. 2023.

[22] W. Tong, Y.-X. Li, X.-Y. Zhao, Q.-Q. Chen, Y.-B. Gao, P. Li, and E. Q. Wu, “fmri-based brain disease diagnosis: A graph network approach,” IEEE. Trans. Med. Robot. Bionics, vol. 5, no. 2, pp. 312–322, 2023.

[23] X. Li, Y. Zhou, N. Dvornek, M. Zhang, S. Gao, J. Zhuang, D. Scheinost, L. H. Staib, P. Ventola, and J. S. Duncan, “BrainGNN: Interpretable Brain Graph Neural Network for fMRI Analysis,” Medical Image Analysis, vol. 74, p. 102233, Dec. 2021.

[24] X. Kan, W. Dai, H. Cui, Z. Zhang, Y. Guo, and C. Yang, “Brain network transformer,” in Advances in Neural Information Processing Systems 35 (NeurIPS 2022), New Orleans, LA, USA, Nov. 2022.

[25] W. Hilal, S. A. Gadsden, and J. Yawney, “Cognitive Dynamic Systems: A Review of Theory, Applications, and Recent Advances,” Proceedings of the IEEE, vol. 111, no. 6, pp. 575–622, Jun. 2023.

[26] J. Yang, S. Gohel, and B. Vachha, “Current methods and new directions in resting state fMRI,” Clinical Imaging, vol. 65, pp. 47–53, Sep. 2020.

[27] M. G. Preti, T. A. Bolton, and D. Van De Ville, “The dynamic functional connectome: State-of-the-art and perspectives,” NeuroImage, vol. 160, pp. 41–54, Oct. 2017.

[28] R. M. Hutchison, T. Womelsdorf, E. A. Allen, P. A. Bandettini, V. D. Calhoun, M. Corbetta, S. Della Penna, J. H. Duyn, G. H. Glover, J. Gonzalez-Castillo, D. A. Handwerker, S. Keilholz, V. Kiviniemi, D. A. Leopold, F. de Pasquale, O. Sporns, M. Walter, and C. Chang, “Dynamic functional connectivity: Promise, issues, and interpretations,” NeuroImage, vol. 80, pp. 360–378, Oct. 2013.

[29] S. Gadgil, Q. Zhao, A. Pfefferbaum, E. V. Sullivan, E. Adeli, and K. M. Pohl, “Spatio-temporal graph convolution for resting-state fMRI analysis,” in Medical Image Computing and Computer Assisted Intervention (MICCAI). Cham: Springer International Publishing, 2020, pp. 528–538.

[30] S. Dahan, L. Z. J. Williams, D. Rueckert, and E. C. Robinson, “Improving phenotype prediction using long-range spatio-temporal dynamics of functional connectivity,” in Machine Learning in Clinical Neuroimaging. Cham: Springer International Publishing, Sep. 2021, pp. 145–154.

[31] L. Yang, C. Chatelain, and S. Adam, “Dynamic Graph Representation Learning With Neural Networks: A Survey,” IEEE Access, vol. 12, pp. 43 460–43 484, 2024.

[32] G. Papanastasiou, N. Dikaios, J. Huang, C. Wang, and G. Yang, “Is Attention all You Need in Medical Image Analysis? A Review,” IEEE Journal of Biomedical and Health Informatics, vol. 28, no. 3, pp. 1398–1411, Mar. 2024.

[33] S. Min, Z. Gao, J. Peng, L. Wang, K. Qin, and B. Fang, “Stgsn — a spatial–temporal graph neural network framework for time-evolving social networks,” Knowledge-Based Systems, vol. 214, p. 106746, Feb. 2021.

[34] S. Rahmani, A. Baghbani, N. Bouguila, and Z. Patterson, “Graph neural networks for intelligent transportation systems: A survey,” IEEE Transactions on Intelligent Transportation Systems, vol. 24, no. 8, pp. 8846–8885, Aug. 2023.

[35] G. Ma, V. A. Vo, T. L. Willke, and N. K. Ahmed, “Memory-augmented graph neural networks: A brain-inspired review,” IEEE Transactions on Artificial Intelligence, vol. 5, no. 5, pp. 2011–2025, May 2024.

[36] J. Guo, C. L. P. Chen, Z. Liu, and X. Yang, “Dynamic Neural Network Structure: A Review for Its Theories and Applications,” IEEE Transactions on Neural Networks and Learning Systems, pp. 1–21, 2024.

[37] Y. Ma, Q. Wang, L. Cao, L. Li, C. Zhang, L. Qiao, and M. Liu, “Multi-Scale Dynamic Graph Learning for Brain Disorder Detection With Functional MRI,” IEEE Transactions on Neural Systems and Rehabilitation Engineering, vol. 31, pp. 3501–3512, 2023.

[38] Q. Zhu, S. Li, X. Meng, Q. Xu, Z. Zhang, W. Shao, and D. Zhang, “Spatio-Temporal Graph Hubness Propagation Model for Dynamic Brain Network Classification,” IEEE Transactions on Medical Imaging, vol. 43, no. 6, pp. 2381–2394, Jun. 2024.

[39] A. Schaefer, R. Kong, E. M. Gordon, T. O. Laumann, X.-N. Zuo, A. J. Holmes, S. B. Eickhoff, and B. T. T. Yeo, “Local-Global Parcellation of the Human Cerebral Cortex from Intrinsic Functional Connectivity MRI,” Cerebral Cortex, vol. 28, no. 9, pp. 3095–3114, Sep. 2018.

[40] P. Ciuciu, J.-B. Poline, G. Marrelec, J. Idier, C. Pallier, and H. Benali, “Unsupervised robust nonparametric estimation of the hemodynamic response function for any fmri experiment,” IEEE Transactions on Medical Imaging, vol. 22, no. 10, pp. 1235–1251, Oct. 2003.

[41] K. Worsley and K. Friston, “Analysis of fmri time-series revisited—again,” NeuroImage, vol. 2, no. 3, pp. 173–181, Sep. 1995.

[42] N. Wiener, Extrapolation, Interpolation, and Smoothing of Stationary Time Series: With Engineering Applications. The MIT Press, Aug. 1949.

[43] R. Killick, P. Fearnhead, and I. A. Eckley, “Optimal detection of changepoints with a linear computational cost,” Journal of the American Statistical Association, vol. 107, pp. 1590–1598, Oct. 2012.

[44] P. Velickovic, G. Cucurull, A. Casanova, A. Romero, P. Lio, and Y. Bengio, “Graph attention networks,” in 6th International Conference on Learning Representations (ICLR). Vancouver, BC, Canada: Open-Review.net, Apr. 2018.

[45] A. Vaswani, N. Shazeer, N. Parmar, J. Uszkoreit, L. Jones, A. N. Gomez, L. Kaiser, and I. Polosukhin, “Attention is all you need,” in Proceedings of the 31st International Conference on Neural Information Processing Systems, ser. NIPS’17. Red Hook, NY, USA: Curran Associates Inc., 2017, pp. 6000–6010.

[46] C. Cortes and V. Vapnik, “Support-vector networks,” Machine Learning, vol. 20, no. 3, pp. 273–297, Sep. 1995.

[47] L. Breiman, “Random Forests,” Machine Learning, vol. 45, no. 1, pp. 5–32, 2001.

[48] D. E. Rumelhart, G. E. Hinton, and R. J. Williams, “Learning representations by back-propagating errors,” Nature, vol. 323, no. 6088, pp. 533–536, Oct. 1986.

[49] T. N. Kipf and M. Welling, “Semi-supervised classification with graph convolutional networks,” in 5th International Conference on Learning Representations, ICLR 2017, Conference Track Proceedings. Toulon, France: OpenReview.net, Apr. 2017.

[50] K. Xu, W. Hu, J. Leskovec, and S. Jegelka, “How powerful are graph neural networks?” in International Conference on Learning Representations (ICLR), New Orleans, LA, USA, May 2019.

